# Characteristics and outcome profile of Hospitalized African COVID-19 patients: The Ethiopian Context

**DOI:** 10.1101/2020.10.27.20220640

**Authors:** Tigist W. Leulseged, Ishmael S. Hassen, Endalkachew H. Maru, Wuletaw C. Zewde, Negat W. Chamiso, Abdi B. Bayisa, Daniel S. Abebe, Birhanu T. Ayele, Kalkidan T. Yegle, Mesay G. Edo, Eyosyas K. Gurara, Dereje D. Damete, Yared A. Tolera

**Author notes:** Divison of Epidemiology and Biostatistics, Faculty of Medicine and Health Sciences, Stellenbosch University, Cape Town, South Africa (Birhanu T. Ayele). Corresponding author: Tigist W. Leulseged, Research development office, Millennium COVID-19 Care Center, Addis Ababa, Ethiopia Department of Internal Medicine, St. Paul’s Hospital Millennium Medical College, Addis Ababa, Ethiopia.

## Abstract

**Background:** The COVID-19 pandemic seems to have a different picture in Africa; the first case was identified in the continent after it has already caused a significant loss to the rest of the world and the reported number of cases and mortality rate has been low. Understanding the characteristics and outcome of the pandemicin the African setup is therefore crucial.

**Aim:** To assess the characteristics and outcome of COVID-19 patients and to identify determinants of the disease outcome among patients admitted to Millennium COVID-19 Care Center in Ethiopia.

**Methods:** A prospective cohort study was conducted among 1345 consecutively admitted RT-PCR confirmed COVID-19 patients from July to September, 2020. Frequency tables, KM plots, median survival times and Log-rank test were used to describe the data and compare survival distribution between groups. Cox proportional hazard survival model was used to identify determinants of time to clinical improvement and the independent variables, where adjusted hazard ratio, P-value and 95% CI for adjusted hazard ratio were used for testing significance and interpretation of results. Binary logistic regression model was used to assess the presence of a statistically significant association between disease outcome and the independent variables, where adjusted odds ratio, P-value and 95% CI for adjusted odds ratio were used for testing significance and interpretation of results

**Results:** Among the study population, 71 (5.3%) died, 72 (5.4%) were transferred and the rest 1202 (89.4%) were clinically improved. The median time to clinical improvement was 14 days. On the multivariable Cox proportional hazard model; temperature (AHR= 1.135, 95% CI= 1.011, 1.274, p-value=0.032), COVID-19 severity (AHR= 0.660, 95% CI= 0.501, 0.869, p-value=0.003), and cough (AHR= 0.705, 95% CI= 0.519, 0.959, p-value=0.026) were found to be significant determinants of time to clinical improvement. On the binary logistic regression, the following factors were found to be significantly associated with disease outcome; SPO2 (AOR= 0.302, 95% CI= 0.193, 0.474, p-value=0.0001), shortness of breath (AOR= 0.354, 95% CI= 0.213, 0.590, p-value=0.0001) and diabetes mellitus (AOR= 0.549, 95% CI= 0.337, 0.894, p-value=0.016).

**Conclusions:** The average duration of time to clinical improvement was 14 days and 89.4 % of the patients achieved clinical improvement. The mortality rate of the studied population is lower than reports from other countries including those in Africa. Having severe COVID-19 disease severity and presenting with cough were found to be associated with delayed clinical improvement of the disease. On the other hand, being hyperthermic is associated with shorter disease duration (faster time to clinical improvement). In addition, lower oxygen saturation and subjective complaint of shortness of breath and being diabetic were associated with unfavorable disease outcome. Therefore, patients with these factors should be followed cautiously for a better outcome.

## INTRODUCTION

Over the past ten months the COVID-19 pandemic has caused a significant loss, both to the human life and the economy, all over the world. The Africa continent, which is already burdened with communicable diseases like HIV and Tuberculosis with limited health care infrastructure and underdeveloped healthcare sector, seemed to be less affected by the pandemic. According to the World Health Organization weekly epidemiological report, as of October 11, 2020 globally there were 37,109,851 confirmed cases. Africa constituted only 3% (1, 227, 719) of the global case. So far, a total of 27,255 deaths were reported in the continent and this constitutes only 3% of the global cumulative death. In Ethiopia, on the same day, there were a total of 83,429 confirmed cases and 1277 deaths ^1^.

Numerous studies are conducted globally to understand the pandemic better, but still studies are scarce in Africa. A recent study conducted in Democratic Republic of Congo, that assessed the characteristics and outcome of COVID-19 patients, showed that mortality rate from COVID-19 was 13.2% and significant predictors were age group, body mass index andhistory of chronic kidney disease^2^.

Different clinical presentation, disease course and outcome have been reported that varies from study to study and place to place implying the need for further study to understand the disease better. Regarding the clinical presentation, symptomatic and asymptomatic presentationshave been reported during the entire disease course. In addition, the type of symptoms could vary manifesting as different body system symptoms with some presenting with what is called atypical presentations for a virus that attacks the respiratory system. The commonly reported symptoms are respiratory and constitutional symptoms^3-9^.

The disease outcome is reported to vary between a complete recovery without any complication, development of one or more systemic complication or death^10-12^. These disease outcomes seemed to be determined by underlying patient characteristics, history of pre-existing medical conditions, the COVID-19 disease severity, disease progression and the development of complication ^11,13-20^.

Due to varying reports from studies conducted in different countries, understanding the disease characteristics and its outcome in the local context is crucial for targeted intervention.

Therefore, in this study we aimed to assess the characteristics and outcome of 1345 RT-PCR confirmed COVID-19 patients and to identify the determinants of the disease outcome among patients admitted to Millennium COVID-19 Care Center in Ethiopia.

## METHODS AND MATERIALS

### Study setting, design and population

An institution based prospective cohort study was conducted at Millennium COVID-19 Care Center (MCCC), a makeshift hospital in Addis Ababa, Ethiopia.

The follow up was made from July, when the center started functioning with full capacity, up to the end of September, 2020.

The source population was all cases of COVID-19 admitted at MCCC with a confirmed diagnosis of COVID-19 using RT-PCR, as reported by a laboratory given mandate to test such patients by the Ministry of Health and who were on follow up from July to September, 2020^21^.

### Sample size Determination and Sampling Technique

All consecutively admitted Severe COVID-19 patients during the three months follow up period were included in the study.

During this interval a total of 1741 COVID-19 patients were admitted to the Center.

### Eligibility criteria

All COVID-19 patients who were on treatment and follow up at the center from July to September, 2020 and with complete follow up data were included.

### Operational Definitions

**Event:** Clinical improvement from COVID-19.

**Censoring:** Includes patients who lost to follow-up, transferred out, died or completed the follow-up period before clinical recovery.

**Time to event or censoring:** time between admission to the Center up to clinical improvement or censoring (in days).

### Data Collection Procedures and Quality Assurance

Data was collected from patients and their medical charts using a pretested interviewer administered questionnaire. Training on the basics of the questionnaire and data collection tool was given for fifteen data collectors (General practitioners) and four supervisors (General practitioner and public health specialist) for two days. The recommended infection prevention and control practice was implemented during the data collection. Data consistency and completeness was checked before an attempt was made to enter the code and analyze the data.

### Data Management and Data Analysis

The collected data was coded and entered into Epi-Info version 7.2.1.0, cleaned and stored and exported into SPSS version 23 for analysis. Data was summarized using frequency tables, Kaplan Meier (KM) plots and median survival times. Survival experience of different groups was compared using KM survival curves. Log-rank test was used to assess significant difference among survival distributions of groups for equality.

To assess the presence of a statistically significant association between the independent variables and time to clinical improvement, multivariable Cox proportional hazard survival model was used. Univariate analysis was performed to calculate an unadjusted hazard ratio (HR) and to screen out potentially independent variables. In the final model; Adjusted HR, P-value and 95% CI for HR were used to test significance and interpretation of results. Variables with p-value ≤ 0.05 were considered as statistically associated with time to clinical improvement in days. The basic assumptions of Cox Proportional Hazard model was tested using log minus log function, where parallel lines indicate proportionality, and the data fitted well.

Similarly, to assess the association between the relevant independent variables and disease outcome, multivariable Binary logistic regression model was used. Univariate analysis was done to screen out independent variables to be used in the multiple Binary Logistic regression model. The adequacy of the final model was assessed using the Hosmer and Lemeshow goodness of fit test and the final model fitted the data well p-value = 0.167). For the Binary Logistic regression, 95% confidence interval for AOR was calculated and variables with p-value ≤ 0.05 were considered as statistically associated with disease outcome.

## RESULT

### Disease outcome, Censoring status and median time to clinical improvement

Among the 1741 patients, 1345 patients with complete medical records were included in the study. Seventy-one (5.3%) died, 72 (5.4%) were transferred and the rest 1202 (89.4%) were discharged improved.

Among the 1345 patients, 1202 (89.4%) achieved clinical improvement and 143 (10.6%) were censored. The overall median time to clinical improvement was 14 days.

### Socio-demographic, comorbid illness and vital sign related variables, censoring status and survival experience

The mean age of the participants was 41.0 ± 18.2 years and 778 (58.6%) were males. The majority of the participants (92.9%) were from Addis Ababa, the capital city of Ethiopia. Only 26 (1.9%) of the participants were health care professionals. From the 557 females, 13 (2.3%) were pregnant.

Around one-third (34.2%) of the patients had a history of one or more spre-existing co-morbid illness. The major co-morbid illness among the participants was hypertension (19.9%), diabetes (13.7%), cardiac disease (4.2%) and asthma (4.1%). Chronic kidney disease, chronic liver disease, neurologic disorder and chronic pulmonary diseases constitute only 2.6 % altogether.

Based on the vital sign recorded on triage, majority had SBP of < 140 (54.7%), DBP of < 90 (72.4%), temperature of > 37.5 °C (52.3%) and oxygen saturation of 93% and above (80.4%).

In all of the variable categories, the number of those who achieved clinical improvement was greater than the censored observations. The proportion of censored observations was relatively larger as age increases, for males, for patients with one or more pre-existing comorbid illness and Khat chewers. On the other, those with a history of drug use had a relatively less censored observation than those with no drug use history.

A statistically significant difference in the time to clinical improvement was observed among patients based on age group, hypertension, cardiac disease, diabetes mellitus and oxygen saturation. Accordingly, the median duration of time to clinical improvement was significantly longer for patients 50 years and older (15 days) compared to those patients younger than 50 years of age (14 days). Having a pre-existing cardiac disease (15 days Vs 14 days) and diabetes mellitus (15 days Vs 14 days) was associated with a delayed clinical improvement compared to patients with no such comorbid illnesses. In addition, delayed clinical improvement was observed among patients whose oxygen saturation was lower than 93% at presentation (16 days Vs 14 days). (**Table 1**)

**Table 1:** Socio–demographic, co-morbid illness and vital sign related variables, censoring status and survival experience among COVID-19 patients (n=1345)

| Variable | Censoring status |  | Total (%) | Median survival time (days) | P-value |
| --- | --- | --- | --- | --- | --- |
|  | No of event (%) | No of censored (%) |  |  |  |
| **Age** |  |  |  |  |  |
| < 30 | 444 (94.8) | 39 (8.1) | 483 (35.9) | 14.0 | **0.0001\*** |
| 30-49 | 421 (93.2) | 36 (7.9) | 457 (34.0) | 14.0 |  |
| $\geq 50$ | 337 (83.9) | 68 (16.8) | 405 (30.1) | 15.0 | |
| **Sex** |  |  |  |  |  |
| Female | 491 (90.3) | 66 (11.8) | 557 (41.4) | 14.0 | 0.961 |
| Male | 711 (91.5) | 77 (9.8) | 778 (58.6) | 14.0 |  |
| **SBP** |  |  |  |  |  |
| < 140 | 681 (91.8) | 79 (10.4) | 706 (54.7) | 14.0 | 0.733 |
| ≥ 140 | 520 (89.9) | 64 (11.0) | 584 (45.3) | 14.0 |  |
| **DBP** |  |  |  |  |  |
| < 90 | 876 (91.9) | 98 (10.1) | 974 (72.4) | 14.0 | 0.195 |
| ≥ 90 | 326 (88.7) | 45 (12.1) | 371 (27.6) | 14.0 |  |
| **Temperature** |  |  |  |  |  |
| ≤ 37.5 °C | 571 (90.8) | 71 (11.1) | 642 (47.7) | 15.0 | 0.095 |
| > 37.5 °C | 631 (91.2) | 72 (10.2) | 703 (52.3) | 14.0 |  |
| **SPO2** |  |  |  |  |  |
| ≥ 93 | 1008 (95.0) | 73 (6.8) | 1081 (80.4) | 14.0 | **0.0001\*** |
| < 93 | 194 (74.6) | 70 (26.5) | 264 (19.6) | 16.0 |  |
| **Preexisting Co-morbid illness** |  |  |  |  |  |
| No | 813 (93.9) | 72 (8.1) | 885 (65.8) | 14.0 | 0.053 |
| Yes | 389 (85.4) | 71 (15.4) | 460 (34.2) | 14.0 |  |
| **Cardiac disease** |  |  |  |  |  |
| No | 1161 (91.8) | 127 (9.9) | 1288 (95.8) | 14.0 | **0.019\*** |
| Yes | 41 (71.9) | 16 (28.1) | 57 (4.2) | 15.0 |  |
| **Diabetes Mellitus** |  |  |  |  |  |
| No | 1054 (92.5) | 107 (9.2) | 1161 (86.3) | 14.0 | **0.005\*** |
| Yes | 148 (81.5) | 36 (19.6) | 184 (13.7) | 15.0 |  |
| **Asthma** |  |  |  |  |  |
| No | 1152 (91.0) | 138 (10.7) | 1290 (95.9) | 14.0 | 0.901 |
| Yes | 50 (90.9) | 5 (9.1) | 55 (4.1) | 14.0 |  |

### Presenting symptom and disease severity related variables, censoring status and survival experience

More than half (52.2%) of the patients were symptomatic at presentation. The majority had cough (43.6%), followed by shortness of breath (21.2%), fatigue (19.6%), fever (18.1%), headache (14.0%), chest pain (12.5%), sorethroat (11.1%), myalgia (10.9%), arthralgia (10.9%), runny nose (6.1%) and nausea/ vomiting (3%).

More than half of the patients (52.3%) were triaged to have mild COVID-19 severity disease and the rest were moderate (24.7%) to severe (23.0%).

According to the log-rank test result, a significantly longer time to clinical improvement was needed among patients with a complaint of fever (15 days Vs 14 days), chest pain (15 days Vs 14 days), nausea/vomiting (16 days Vs 14 days), fatigue (15 days Vs 14 days), shortness of breath (15 days Vs 14 days) and headache (15 days Vs 14 days). In addition, time to clinical improvement seemed to differ based on disease severity, with sever disease associated with a prolonged time compared to those patients with mild and moderate disease (16 days Vs 14 days).(**Table 2**)

**Table 2:** Presenting symptom related variables censoring status and survival experience among COVID-19 patients (n=1345)

| Variables | Censoring status |  | Total (%) | Median survival time (days) | P-value |
| --- | --- | --- | --- | --- | --- |
|  | No of event (%) | No of censored (%) |  |  |  |
| **Presence of symptom** |  |  |  |  |  |
| Asymptomatic | 601 (36.3) | 42 (6.5) | 643 (47.8) | 14.0 | **0.0001\*** |
| Symptomatic | 601 (86.2) | 101 (14.4) | 702 (52.2) | 15.0 |  |
| **Fever** |  |  |  |  |  |
| No | 992 (91.7) | 110 (10.0) | 1102 (81.9) | 14.0 | **0.0001\*** |
| Yes | 210 (87.7) | 33 (13.6) | 243 (18.1) | 15.0 |  |
| **Cough** |  |  |  |  |  |
| No | 707 (95.5) | 52 (6.9) | 759 (56.4) | 14.0 | **0.0001\*** |
| Yes | 495 (85.2) | 91 (15.5) | 586 (43.6) | 15.0 |  |
| **Nausea/ vomiting** |  |  |  |  |  |
| No | 1166 (91.1) | 138 (10.6) | 1304 (97.4) | 14.0 | **0.026\*** |
| Yes | 36 (97.8) | 5 (12.2) | 41 (3.0) | 16.0 |  |
| **Sore throat** |  |  |  |  |  |
| No | 1065 (90.8) | 131 (11.0) | 1196 (88.9) | 14.0 | 0.340 |
| Yes | 137 (92.6) | 12 (8.1) | 149 (11.1) | 14.0 |  |
| **Runny nose** |  |  |  |  |  |
| No | 1125 (90.8) | 138 (10.9) | 1263 (93.9) | 14.0 | 0.226 |
| Yes | 77 (93.9) | 5 (6.1) | 82 (6.1) | 14.0 |  |
| **Chest pain** |  |  |  |  |  |
| No | 1058 (91.7) | 119 (10.1) | 1177 (87.5) | 14.0 | **0.0001\*** |
| Yes | 144 (86.3) | 24 (14.3) | 168 (12.5) | 15.0 |  |
| **Myalgia** |  |  |  |  |  |
| No | 1070 (91.1) | 128 (10.7) | 1198 (89.1) | 14.0 | 0.302 |
| Yes | 132 (91.5) | 15 (10.2) | 147 (10.9) | 15.0 |  |
| **Arthralgia** |  |  |  |  |  |
| No | 1074 (91.3) | 125 (10.4) | 1199 (89.1) | 14.0 | 0.160 |
| Yes | 128 (88.4) | 18 (18.0) | 146 (10.9) | 15.0 |  |
| **Fatigue** |  |  |  |  |  |
| No | 984 (93.0) | 97 (9.0) | 1081 (80.4) | 14.0 | **0.0001\*** |
| Yes | 218 (83.0) | 46 (17.4) | 264 (19.6) | 15.0 |  |
| **SOB** |  |  |  |  |  |
| No | 986 (94.9) | 74 (7.0) | 1060 (78.8) | 14.0 | **0.0001** |
| Yes | 216 (76.5) | 69 (24.2) | 285 (21.2) | 15.0 |  |
| **Headache** |  |  |  |  |  |
| No | 1041 (91.9) | 115 (9.9) | 1156 (85.9) | 14.0 | **0.012\*** |
| Yes | 161 (84.7) | 28 (14.8) | 189 (14.1) | 15.0 |  |
| **COVID-19 Severity** |  |  |  |  |  |
| Mild | 656 (96.2) | 47 (6.7) | 703 (52.3) | 14.0 | **0.0001\*** |
| Moderate | 316 (95.5) | 16 (4.8) | 332 (24.7) | 14.0 |  |
| Severe | 230 (74.5) | 80 (25.8) | 310 (23.0) | 16.0 |  |

The Kaplan Meir (KM) survival function graphs also showed that those with mild disease and no symptoms at presentation have a favorable survival experience (time to clinical improvement) compared to those with more severe disease category and symptomatic diseases. (**Figure 2**)

**Figure 1:**
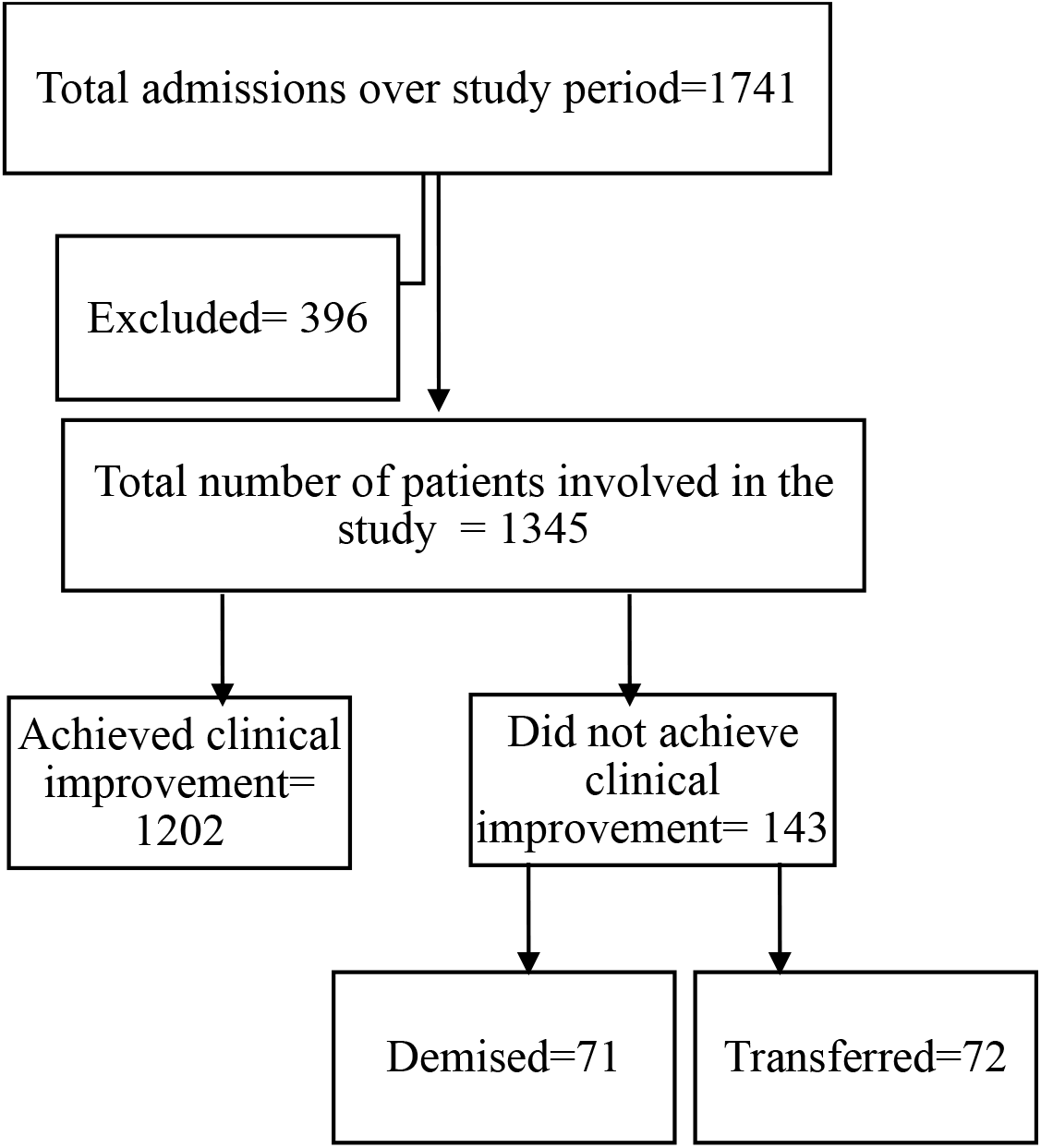
Flow chart showing the disposition of study participants in the final analysis

**Figure 2:**
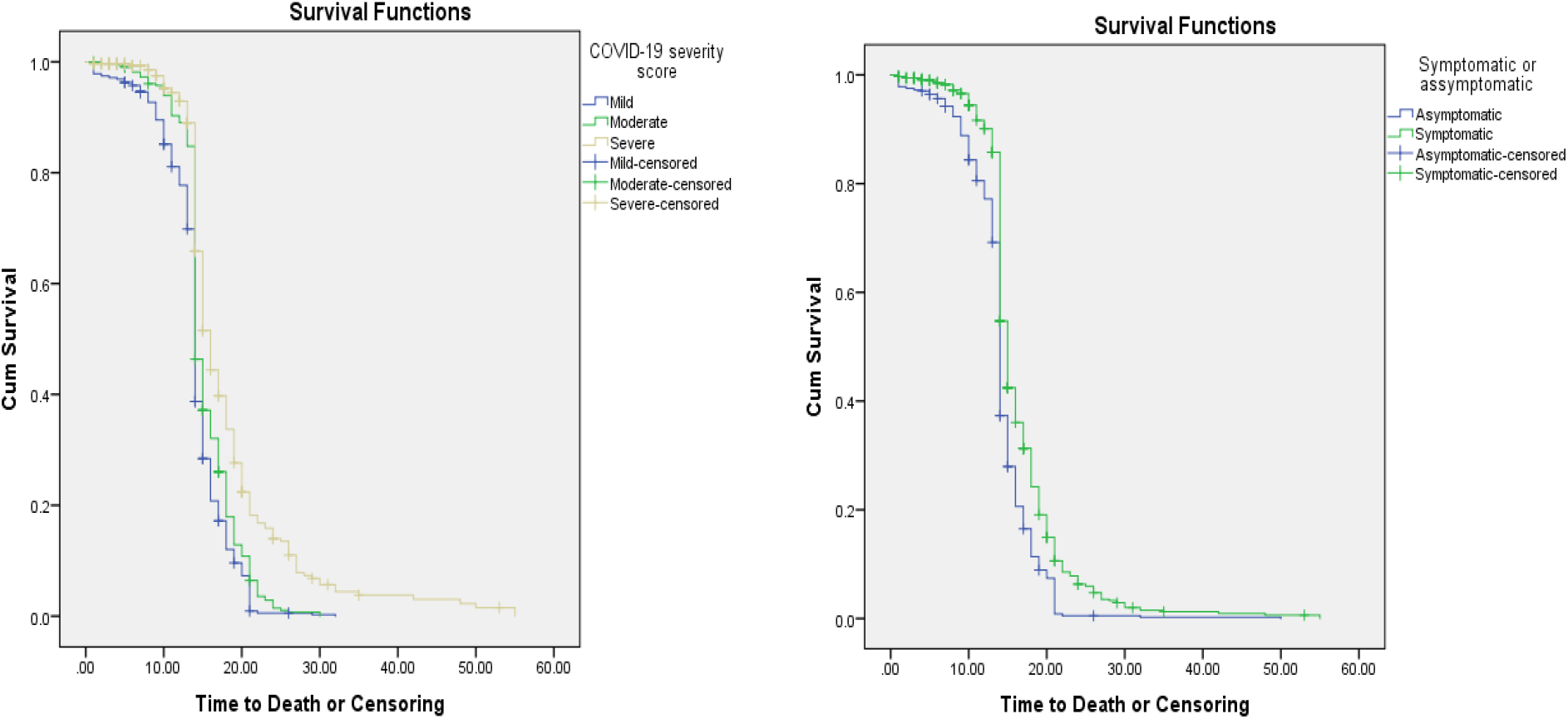
Survival functions for COVID severity and Presence of symptom by time

### Factors affecting time to clinical improvement from COVID-19

Crude analysis of each independent variable with the time to clinical improvement was run at 25% level of significance. From univariate analysis; age group, SPO2, temperature, COVID-19 severity, cough, chest pain, arthralgia, fatigue, headache, shortness of breath,hypertension and diabeteswere significantly associated with time to clinical improvement among COVID-19 patients.

However; only temperature, COVID-19 severity and cough were found to be significantly associated with time to clinical improvement in the multivariable Cox proportional hazard model at 5% level of significance.The proportional hazards assumption of the model was tested using the Log-minus-Log function on SPSS version 23 software. The plots show a reasonable fit to the assumption with Parallel lines between groups indicate proportionality ^22^.

Accordingly, after adjusting for other covariates, the rate of achieving clinical improvement among patients with temperature of 37.5 °C and above was 1.135 times than patients with a temperature range of37.5 °C and lower (AHR= 1.135, 95% CI= 1.011, 1.274, p-value=0.032). This shows that hyperthermic patients improve from COVID-19 in a significantly shorter duration.

COVID-19 disease severity at admission was also found to be a significant determinant of time to clinical improvement. The rate of achieving clinical improvement among patients with severe COVID-19 disease at presentation was 34% lower than patients who presented with mild disease (AHR= 0.660, 95% CI= 0.501, 0.869, p-value=0.003). On the other hand, there was no significant difference between moderate and mild cases in the time to clinical improvement. That means, having severe disease is associated with delayed clinical improvement.

Presenting with a complaint of cough at admission was associated with a 24.1% lower rate of achieving clinical improvement compared to those patients with no such complaint on admission(AHR= 0.705, 95% CI= 0.519, 0.959, p-value=0.026) showing that having cough resulted in delayed clinical improvement (**Table 3**)

**Table 3:**
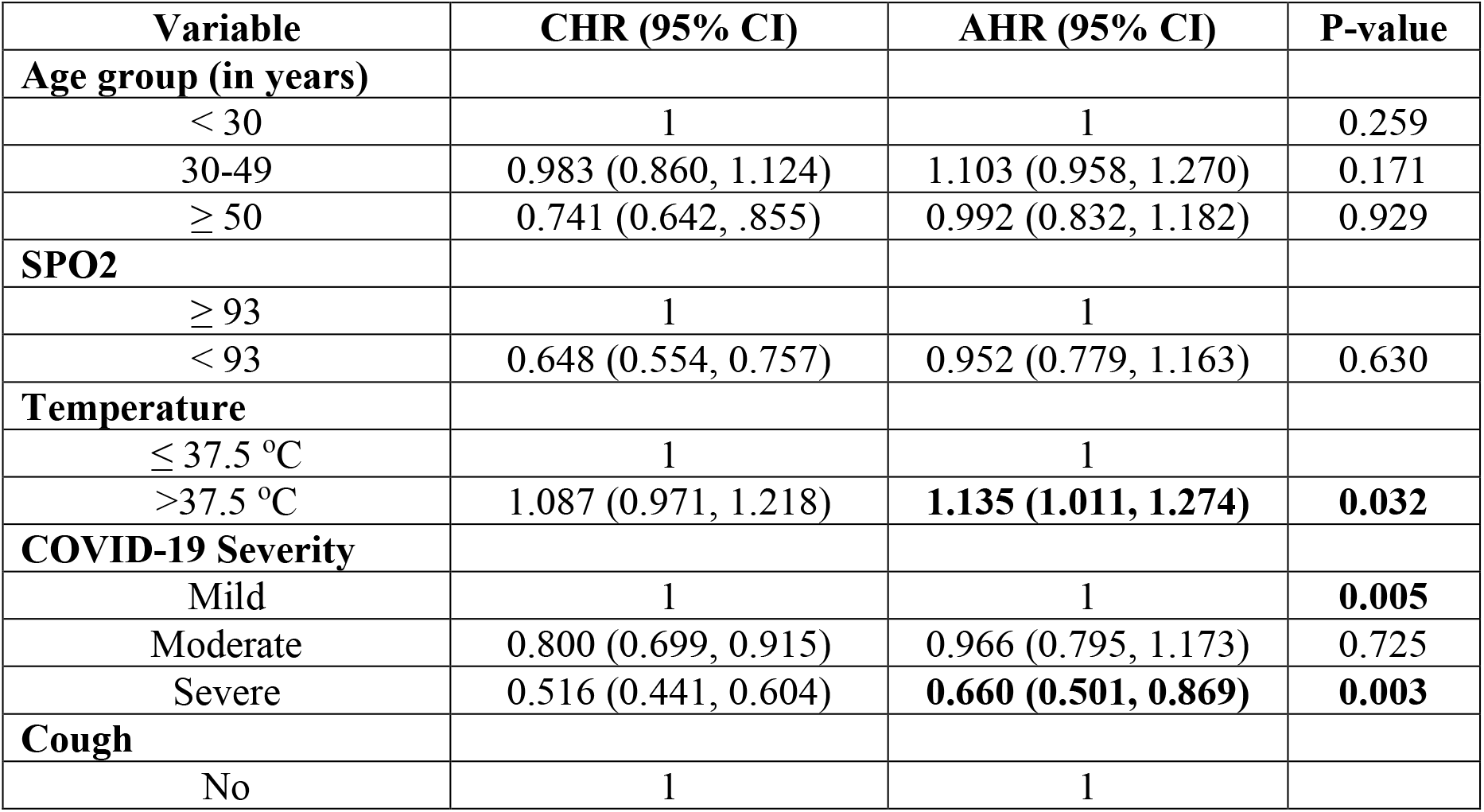
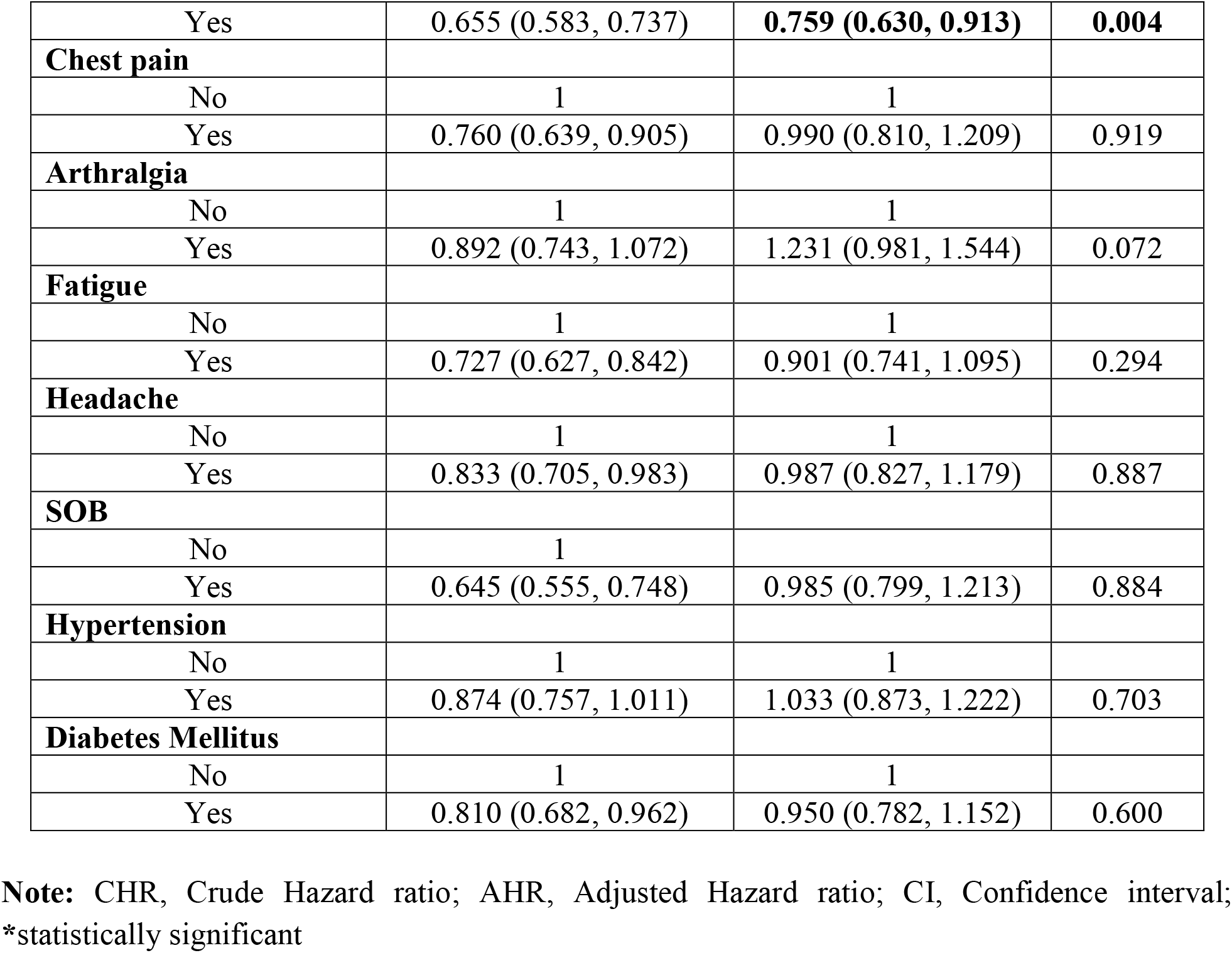
Result of Multivariable Cox proportional hazard model among COVID-19 patients (n=1345)

### Factors affecting disease outcome

Univariate analysis at 25% level of significance was conducted and age group, sex, SPO2, fever, cough, chest pain, arthralgia, fatigue, shortness of breath, headache, hypertension and diabetes mellitus were found to be significantly associated with the clinical improvement from COVID-19 disease.

On the multivariable binary logistic regression, after adjusting for other covariates, SPO2, shortness of breath and diabetes mellitus were found to be significantly associated with achievement of clinical improvement at 5% level of significance.

Both the subjective complaint of shortness of breath and the objectively measured oxygen saturation level were found to be significant determinants of achievement of clinical improvement. Accordingly, after adjusting for other covariates, the odds of achieving clinical improvement among patients with SPO2 of lower than 93% and a subjective complaint of shortness of breath were 69.8% and 64.6% lower than patients with SPO2 of 93% and above and with no shortness of breath, respectively (AOR= 0.302, 95% CI= 0.193, 0.474, p-value=0.0001 for SPO2 and AOR= 0.354, 95% CI= 0.213, 0.590, p-value=0.0001 for shortness of breath).

Having diabetes mellitus was associated with a 45.1% lower odds of achieving clinical improvement compared to those with no such comorbid illness (AOR= 0.549, 95% CI= 0.337, 0.894, p-value=0.016). (**Table 4**)

**Table 4:** Results for the final multivariable binary logistic regression model among COVID-19 patients (n=1345)

| Variables | Outcome |  | COR(95% CI) | AOR(95% CI) | p-value |
| --- | --- | --- | --- | --- | --- |
|  | Clinically improved | Dead and transferred |  |  |  |
| **Age group (in years)** |  |  |  |  |  |
| < 30 | 444 (91.9) | 39 (8.1) |  |  | 0.187 |
| 30-49 | 421 (92.1) | 36 (7.9) | 1.027 (0.641, 1.647) | 1.622 (0.958, 2.749) | 0.072 |
| ≥ 50 | 337 (83.2) | 68 (16.8) | 0.435 (0.286, 0.661) | 1.254 (0.711, 2.213) | 0.435 |
| **Sex** |  |  |  |  |  |
| Female | 491 (88.2) | 66 (11.8) |  |  |  |
| Male | 711 (90.2) | 77 (9.8) | 1.241 (0.876, 1.758) | 1.445 (0.981, 2.129) | 0.063 |
| **SPO2** |  |  |  |  |  |
| No | 1008 (93.2) | 73 (6.8) |  |  |  |
| Yes | 194 (73.5) | 70 (26.5) | 0.201 (0.140, 0.288) | **0.302 (0.193, 0.474)** | **0.0001** |
| **Fever** |  |  |  |  |  |
| No | 992 (90.0) | 110 (10.0) |  |  |  |
| Yes | 210 (86.4) | 33 (13.6) | 0.706 (0.465, 1.070) | 1.385 (0.822, 2.334) | 0.220 |
| **Cough** |  |  |  |  |  |
| No | 707 (93.1) | 52 (6.9) |  |  |  |
| Yes | 495 (84.5) | 91 (15.5) | 0.400 (0.279, 0.573) | 0.720 (0.449, 1.156) | 0.174 |
| **Chest pain** |  |  |  |  |  |
| No | 1058 (89.9) | 119 (10.1) |  |  |  |
| Yes | 144 (85.7) | 24 (14.3) | 0.675 (0.421, 1.082) | 1.325 (0.756, 2.322) | 0.325 |
| **Arthralgia** |  |  |  |  |  |
| No | 1074 (89.6) | 125 (10.4) |  |  |  |
| Yes | 128 (87.7) | 18 (12.3) | 0.828 (0.489, 1.402) | 1.508 (0.801, 2.839) | 0.204 |
| **Fatigue** |  |  |  |  |  |
| No | 984 (91.0) | 97 (9.0) |  |  |  |
| Yes | 218 (82.6) | 46 (17.4) | 0.467 (0.319, 0.683) | 1.034 (0.617, 1.732) | 0.900 |
| **SOB** |  |  |  |  |  |
| No | 986 (93.0) | 74 (7.0) |  |  |  |
| Yes | 216 (75.8) | 69 (24.2) | 0.235 (0.164, 0.337) | **0.354 (0.213, 0.590)** | **0.0001** |
| **Headache** |  |  |  |  |  |
| No | 1041 (90.1) | 115 (9.9) |  |  |  |
| Yes | 161 (85.2) | 28 (14.8) | 0.635 (0.407, 0.991) | 0.830 (0.496, 1.388) | 0.477 |
| **Hypertension** |  |  |  |  |  |
| No | 973 (90.3) | 105 (9.7) |  |  |  |
| Yes | 229 (85.8) | 38 (14.2) | 0.650 (0.437, 0.968) | 1.090 (0.664, 1.729) | 0.733 |
| **Diabetes mellitus** |  |  |  |  |  |
| No | 1054 (90.8) | 107 (9.2) |  |  |  |
| Yes | 148 (80.4) | 36 (19.6) | 2.396 (1.582, 3.628) | **0.549 (0.337, 0.894)** | **0.016** |

## DISCUSSION

In this study, we assessed the characteristics and outcome profile of 1345 RT-PCR confirmed COVID-19 patients admitted to Millennium COVID-19 Care Center in Ethiopia from July to September, 2020. There is scarcity of research in African COVID-19 patients that describes the characteristics and outcome of patients and what affects it. Understanding these factors will guide policy makers in making evidence based decision. Among the study population, 71 (5.3%) died, 72 (5.4%) were transferred and the rest 1202 (89.4%) were discharged improved. The median time to clinical improvement was 14 days. The death rate is low compared to studies from other countries including a study from Democratic republic of Congo, where a death rate of 13.2% was reported ^2^. Studies conducted in Italy and New York also reported a death rate of up to 23% ^12,20^.

On the log-rank test, less favorable survival experience was observed among patients who were 50 years and older, those with cardiac illness, diabetes mellitus, oxygen saturation of less than 93%, those with severe disease and those who are symptomatic particularly those who presented with fever, cough, chest pain, fatigue, shortness of breath, headache and nausea/ vomiting.

On the multivariable Cox proportional hazard model; temperature, COVID-19 severity and cough were found to be significant determinants of time to clinical improvement.

Accordingly, after adjusting for other covariates, the rate of achieving clinical improvement among patients with body temperature of above 37.5 °C was 1.135 times than patients with temperature of 37.5 °C and lower. This shows that those patients with an objectively recorded hyperthermia have a favorable survival experience. In another case control study conducted in our center as well subjective report of fever by patients was found to be associated with a favorable outcome of being discharged alive. On the other hand, another study in the Center shows that having fever is associated with development of a more severe disease. This could be because having fever is an implication of competent body defense system that can lead to better outcome from the disease even though it could be associated with development of a more severe disease category^23,24^.

COVID-19 disease severity at admission was also found to be a significant determinant of time to clinical improvement. The rate of achieving clinical improvement among patients with severe COVID-19 disease at presentation was 34% lower than patients who presented with mild disease. On the other hand, there was no significant difference between moderate and mild cases in the time to clinical improvement. Severe patients are those patients with symptoms and signs that could lead to intensive care admission and that were also found to be significant predictors of unfavorable disease outcome in this study as well.

Presenting with a complaint of cough at admission was associated with a 24.1% lower rate of achieving clinical improvement compared to those patients with no such complaint on admission. This could be because those patients with cough were in a more severe disease category, which was also found to be a significant determinant of time to clinical improvement. Another study conducted in the Center on determinants of time to convalescence also showed that having symptomatic disease in general is associated with a delayed biochemical recovery^25^.

On the binary logistic regression at 5% level of significance, the following factors were found to be significantly associated with disease outcome;SPO2, shortness of breath, and diabetes mellitus.

Both the subjective complaint of shortness of breath and the objectively measured oxygen saturation level were found to be significant determinants of disease outcome. Accordingly, after adjusting for other covariates, the odds of achieving clinical improvement among patients with SPO2 of lower than 93% and a subjective complaint of shortness of breath were 69.8% and 64.6% lower than patients with SPO2 of 93% and above and with no shortness of breath, respectively. This could be because shortness of breath and decreased SPO2 are manifestations of diseased lung with diminished capacity, which can be explained also by the high affinity of the SARS-COV-2 virus to attack the lung. Shortness of breath is also found to be associated with increased odds of death and also prolonged oxygen requirement among severe COVID-19 patients in other studies conducted in our Center ^24,26^.

Having diabetes mellitus was associated with a 45.1% lower odds of achieving clinical improvement compared to those with no such comorbid illness. The effect of concomitant comorbid illness on disease progression is also reported to be the same in other studies ^27-30^. Patients with diabetes mellitus were also found to increased likelihood of having death outcome, developing symptomatic disease and more severe disease category in studies conducted in our center ^23,24,31^.

On the other hand, age which was found to be a significant determinant in other studies didn’t show any significant difference in this study ^2,20^.

## CONCLUSION

The average duration of time to clinical improvement was 14 days. The mortality rate of the studied population was 5.3%, this is lower than reports from other countries including Africa.

Having severe COVID-19 disease severity and presenting with cough were found to be associated with delayed clinical improvement from the disease. On the other hand, being hyperthermic is associated with shorter disease duration (faster time to clinical improvement). In addition, lower oxygen saturation and subjective complaint of shortness of breath and being diabetic were associated with unfavorable disease outcome.

Therefore, cautious management of these patients is mandatory for a better management outcome.

## Data Availability

All relevant data are available upon reasonable request

## Declaration

### Ethical Considerations

The study was conducted after obtaining ethical clearance from St. Paul’s Hospital Millennium Medical College Institutional Review Board. Written informed consent was obtained from the participants. The study had no risk/negative consequence on those who participated in the study. Medical record numbers were used for data collection and personal identifiers were not used in the research report. Access to the collected information was limited to the principal investigator and confidentiality was maintained throughout the project.

### Authors Contribution

conceived the study. designed the study, revised data extraction sheet, performed statistical analysis, and drafted the initial manuscript. All authors contributed to the conception of the study and obtained patient data. All authors undertook review and interpretation of the data. All authors revised the manuscript and approved the final version.

## Acknowledgment

The authors would like to thank St. Paul’s Hospital Millennium Medical College for facilitating the research work.

## Availability of data and materials

All relevant data are available upon reasonable request.

## REFERENCES

1. World Health Organization. Weekly COVID-19 Epidemiological Update. 11 October, 2020.

2. Nachega JB, Ishoso DK, Otokoye JO, et al. Clinical Characteristics and Outcomes of Patients Hospitalized for COVID-19 in Africa: Early Insights from the Democratic Republic of the Congo.. Am J Trop Med Hyg. 2020 Oct 2.

3. Avula A, Nalleballe K, Narula N, et al. COVID-19 presenting as stroke. Brain Behav Immun. 2020 Apr 2 8.

4. Taxonera C, Sagastagoitia I, Alba C, Mañas N, Olivares D, Rey E. 2019 Novel Coronavirus Disease (COVID-19) in patients with Inflammatory Bowel Diseases. Aliment Pharmacol Ther 2020 May 02.

5. Aggarwal S, Garcia-Telles N, Aggarwal G, al e. Clinical features, laboratory characteristics, and outcomes of patients hospitalized with coronavirus disease 2019 (COVID-19): Early report from the United States. Dia g n o sis (B e rl). 2020 May 26;7(2):91–96.

6. Hu Y, Sun J, Dai Z, et al. Prevalence and severity of corona virus disease 2019 (COVID- 19): A systematic review and meta-analysis. J Clin Virol. 2020 April 14.

7. Kim G, Kim M, Ra S, et al. Clinical characteristics of asymptomatic and symptomatic patients with mild COVID-19. Clin Microbiol Infect. 2020 April 30.

8. Lechien J, Chiesa-Estomba C, Place S, et al. Clinical and Epidemiological Characteristics of 1,420 European Patients with mild-to-moderate Coronavirus Disease 2019. J Intern Med. 2020 April 30.

9. Xu T, Huang R, Zhu L, et al. Epidemiological and clinical features of asymptomatic patients with SARS-CoV-2 infection. J Med Virol 2020 April 28.

10. Bellosta R, Luzzani L, Natalini G, et al. Acute limb ischemia in patients with COVID-19 pneumonia. J Vasc Surg. 2020 Apr 2 8.

11. Pei G, Zhang Z, Peng J, et al. Renal Involvement and Early Prognosis in Patients with COVID-19 Pneumonia. J Am Soc Nephrol. 2020 Apr 2 8.

12. Richardson S, Hirsch J, Narasimhan M, et al. Presenting Characteristics, Comorbidities, and Outcomes Among 5700 Patients Hospitalized With COVID-19 in the New York City Area. JAMA. 2020 April 22.

13. Bezzio C, Saibeni S, Variola A, et al. Outcomes of COVID-19 in 79 patients with IBD in Italy: an IG-IBD study. Gut. 2020 Apr 30.

14. Mi B, Chen L, Xiong Y, Xue H, Zhou W, Liu G. Characteristics and Early Prognosis of COVID-19 Infection in Fracture Patients. Bone Joint Surg Am. 2020 Apr 0 1

15. Sun L, Shen L, Fan J, et al. Clinical Features of Patients with Coronavirus Disease 2019 (COVID-19) from a Designated Hospital in Beijing, China. J Med Virol 2020 May 05.

16. Wang F, Yang Y, Dong K, et al. CLINICAL CHARACTERISTICS OF 28 PATIENTS WITH DIABETES AND COVID-19 IN WUHAN, CHINA. Endocr Pract. 2020 May 0 1.

17. Yan Y, Yang Y, Wang F, et al. Clinical characteristics and outcomes of patients with severe covid-19 with diabetes BMJ Open Diabetes Res Care. 2020 April;8(1).

18. Yang F, Shi S, Zhu J, Shi J, Dai K, Chen X. Clinical characteristics and outcomes of cancer patients with COVID-19. J Med Virol 2020 May 05.

19. Yang Y, Shen C, Li J, et al. Plasma IP-10 and MCP-3 levels are highly associated with disease severity and predict the progression of COVID-19. J Allergy Clin Immunol 2020 Apr 2 9.

20. Zangrillo A, Beretta L, Scandroglio A, et al. Characteristics, treatment, outcomes and cause of death of invasively ventilated patients with COVID-19 ARDS in Milan, Italy. Crit Care Resusc 2020 April 23.

21. Ethiopian Federal Ministry of Health. Covid19 Management Handbook. 2020.

22. David Hosmer and Lemeshow. Applied survival analysis. 2nd edition ed 2008.

23. Leulseged TW, Kg A, Hassen IS, et al. COVID-19 Disease Severity and Determinants among Ethiopian Patients: A study of the Millennium COVID-19 Care Center. medRxiv 2020.10.09.20209999. 2020.

24. Maru EH, Leulseged TW, Hassen IS, et al. Predictors of Death in Severe COVID-19 Patients at Millennium COVID-19 Care Center in Ethiopia: A Case-Control Study. medRxiv. 2020:2020.2010.2007.20205575.

25. Leulseged TW, Hassen IS, Maru EH, et al. Determinants of Time to Convalescence among COVID-19 Patients at Millennium COVID-19 Care Center in Ethiopia: A prospective cohort study. medRxiv. 2020:2020.2010.2007.20208413.

26. Leulseged TW, Hassen IS, Edo MG, et al. Duration of Oxygen Requirement and Predictors in Severe COVID-19 Patients in Ethiopia: A Survival Analysis. medRxiv. 2020:2020.2010.2008.20209122.

27. Du R-H, Liang L-R, Yang C-Q, et al. Predictors of mortality for patients with COVID-19 pneumonia caused by SARSCoV-2: a prospective cohort study. Eur Respir J. 2020;2020(55: 2000524).

28. Gupta S, Hayek SS, Wang W, et al. Factors Associated With Death in Critically Ill Patients With Coronavirus Disease 2019 in the US. JAMA Intern Med. 2020; e203596.

29. Williamson E.J., Walker A.J., Bhaskaran K, et al. Factors associated with COVID-19- related death using OpenSAFELY. Nature. 584:430–436 (2020).

30. Xiaochen Li, Shuyun Xu, Muqing Yu, et al. Risk factors for severity and mortality in dult COVID-19 inpatients in Wuhan. J ALLERGY CLIN IMMUNOL. 2020;146:110–118.

31. Leulseged TW, Alemahu DG, Hassen IS, et al. Determinants of Developing Symptomatic Disease in Ethiopian COVID-19 Patients. medRxiv 2020.10.09.20209734.

